# OtoXNet - Automated Identification of Eardrum Diseases from Otoscope Videos: A Deep Learning Study for Video-representing Images

**DOI:** 10.1101/2021.08.05.21261672

**Authors:** Hamidullah Binol, M. Khalid Khan Niazi, Charles Elmaraghy, Aaron C. Moberly, Metin N. Gurcan

**Affiliations:** Center for Biomedical Informatics, Wake Forest School of Medicine, Winston-Salem, NC, USA; Department of Otolaryngology, Ohio State University, OH, USA

**Keywords:** Eardrum abnormalities, Computer-assisted Diagnosis, Convolutional Neural Networks, Otoscope, Video classification

## Abstract

**Background:** The lack of an objective method to evaluate the eardrum is a critical barrier to an accurate diagnosis. Eardrum images are classified into normal or abnormal categories with machine learning techniques. If the input is an otoscopy video, a traditional approach requires great effort and expertise to manually determine the representative frame(s).

**Methods:** In this paper, we propose a novel deep learning-based method, called OtoXNet, which automatically learns features for eardrum classification from otoscope video clips. We utilized multiple composite image generation methods to construct a highly representative version of otoscopy videos to diagnose three major eardrum diseases, i.e., otitis media with effusion, eardrum perforation, and tympanosclerosis versus normal (healthy). We compared the performance of OtoXNet against methods with that either use a single composite image or a keyframe selected by an experienced human. Our dataset consists of 394 otoscopy videos from 312 patients and 765 composite images before augmentation.

**Results:** OtoXNet with multiple composite images achieved 84.8% class-weighted accuracy with 3.8% standard deviation, whereas with the human-selected keyframes and single composite images, the accuracies were respectively, 81.8% ± 5.0% and 80.1% ± 4.8% on multi-class eardrum video classification task using an 8-fold cross-validation scheme. A paired t-test shows that there is a statistically significant difference (p-value of 1.3 × 10^−2^) between the performance values of OtoXNet (multiple composite images) and the human-selected keyframes. Contrarily, the difference in means of keyframe and single composites was not significant (p = 5.49 × 10^−1^). OtoXNet surpasses the baseline approaches in qualitative results.

**Conclusion:** The use of multiple composite images in analyzing eardrum abnormalities is advantageous compared to using single composite images or manual keyframe selection.

## 1. Introduction

Diseases of the middle ear and eardrum (tympanic membrane, TM) are the most commonly treated childhood disorders. Treatment is fueled by concern for complications and effects of hearing loss on children’s cognitive and language development (Physicians 2004). Because ear diseases are so common, a major problem is the over-diagnosis and over-treatment of these diseases. Over 8 million unnecessary antibiotics are prescribed annually, contributing to the rise of antibiotic-resistant bacteria (Pelton 1998), and creating the largest number of pediatric medication-related adverse events (Kaleida and Stool 1992). Many children with inaccurate ear diagnoses are referred to Ear, Nose, and Throat (ENT) physicians for surgical placement of ear tubes, and up to 70% of these cases are not indicated (Lieberthal et al. 2013).

The problem of over-diagnosis and over-treatment of ear pathologies is directly related to the diagnoses’ subjective nature when examining the ear using a standard hand-held otoscope, which typically provides a clinician with a brief glimpse of the eardrum. The lack of an objective method to evaluate the eardrum is a critical barrier to an accurate diagnosis. For example, clinicians’ current diagnostic accuracy for ear pathologies is low: in a study of 514 pediatricians and 188 Ear, Nose, and Throat (ENT) physicians viewing videotaped otoscopic exams, diagnostic accuracy rates were only 50% and 73%, respectively (Sorrento and Pichichero 2001). Clearly, new technologies are needed to assist in a more accurate, consistent, and objective diagnosis of ear diseases.

Recent studies have applied computer-assisted image analysis (CAIA) approaches to diagnose common ear pathologies (Mironică, Vertan, and Gheorghe 2011; Kuruvilla et al. 2013; Shie et al. 2014). Mironica *et al*. (Mironică, Vertan, and Gheorghe 2011) have reported an otitis media detection accuracy of 73.1% by applying artificial neural networks and using global features such as color descriptors on a dataset of 186 otoscopic images. They have concluded that color information alone is not sufficient for accurate classification. Kuruvilla *et al*. (Kuruvilla et al. 2013) conducted research to automatically differentiate eardrums with acute otitis media (AOM) from those having no effusion and otitis media with effusion (hereafter referred to as “effusion”) (Myburgh et al. 2016). Those authors stated that their simple and concise 8-feature otitis media vocabulary (visual features used by otoscopists) is effective (with 89.9% accuracy on their dataset) on the problem of automated classification of diagnostic categories of otitis media by highlighting the capability of using physiologically meaningful features. On the other hand, our implementation of the handcrafted features described by Kuruvilla et al. was not able to reach satisfactory results on our dataset compare to the results presented in their paper, since their technique heavily depends on manual and tailored feature selection. Lee et al. utilized a convolutional neural network (CNN) (Lee, Choi, and Chung 2019) to classify eardrums into two categories (normal and perforation) and also to recognize the laterality (right versus left) of the eardrums. They have reached 91.0% accuracy of detecting an eardrum perforation and showed that CNNs can be an effective mechanism for identifying eardrum abnormalities. Shie *et al*. (Shie et al. 2014) proposed an Adaboost-based classifier employing various color, geometric, and texture features to detect otitis media, achieving an accuracy of 88.1%. A recent study (Tran et al. 2018) has reported a 91.4% classification accuracy in distinguishing AOM from effusion using multiple image features such as color and shape with multitask joint sparse representation-based classification algorithm.

Our study is motivated by two considerations: 1) the desire to study multiple ear diseases other than AOM, 2) the utility of videos instead of single images to capture more information necessary for computerized analysis. For the first consideration, the main goal of prior studies was to label an eardrum as normal versus a form of otitis media. Most studies reported only moderate to high classification accuracies between normal versus otitis media. However, many other forms of ear pathology are clinically important because they contribute to hearing loss. For instance, perforations of the eardrum, particularly in children, may result in hearing loss that puts them at risk for delays in language development. Moreover, other high-risk pathologies that are subtle on otoscopy can lead to severe co-morbidities and complications. For example, cholesteatoma, a skin cyst within the middle ear, is often missed by primary care providers but can cause severe hearing loss, dizziness, facial weakness, and even intracranial abscess (Rosito et al. 2016). Similarly, relatively mild eardrum retractions are important abnormalities that need to be monitored because they can develop into cholesteatoma or cause hearing loss themselves. In our prior research, we took advantage of both deep learning techniques (Senaras et al. 2018; Binol, Moberly, et al. 2020a) and approaches that used clinically motivated eardrum features (Senaras et al. 2017) to detect a wide range of eardrum pathologies.

Previously, we developed an otoscopy image analysis algorithm, referred to as “AutoScope” (Senaras et al. 2017), to classify eardrum abnormalities as “normal” versus “abnormal.” First, we developed a preprocessing step to reduce camera-specific problems, detect the region of interest in the image, and prepare the image for further analysis. Subsequently, we designed a new set of clinically motivated eardrum features and evaluated the potential use of visual MPEG-7 descriptors for the task of eardrum image classification. AutoScope was able to classify the eardrum images as normal or abnormal with 84.6% accuracy. In a follow-up study, we began applying deep learning approaches to help with feature extraction in the machine learning workflow (Senaras et al. 2018). We developed an approach to report the eardrum condition of the eardrum as normal or abnormal by an ensemble of two different deep learning architectures, which are Inception-v3 (Szegedy et al. 2016) and a convolutional auto-encoder. The individual classification accuracies of the networks were calculated as 84.4% and 82.6%, respectively. Only 32% of the errors of both networks were the same, making it possible to combine the two approaches (i.e., decision fusion) to achieve better classification accuracy.

In another study, we assessed a decision fusion mechanism to combine predictions obtained from digital otoscopy images and tympanometry data for the detection of eardrum abnormalities. Our database consisted of 73 tympanometry records along with digital otoscopy videos. After obtaining predictions from each of three different classifiers (i.e., random forest on the raw tympanometry data, Inception-ResNet-v2 on the eardrum images, clinical decision tree), we performed a majority voting-based decision fusion technique to reach the final decision. Experimental results showed that the proposed decision fusion method improved the classification accuracy, positive predictive value, and negative predictive value in comparison with the single classifiers. The accuracies were 64.4% for the normal range of the tympanogram values, 76.7% for the computerized analysis of tympanometry data, and 74.0% for the eardrum image analysis, while our decision fusion methodology increased the classification accuracy to 84.9% using leave-one-patient-out cross-validation.

We also designed a deep learning-based content-based image retrieval (CBIR) system, called OtoMatch (Camalan et al. 2020), for differentiating normal eardrum, effusion, and tympanostomy tube conditions. On 10-fold cross-validation, the proposed method resulted in an average accuracy of 80.6% (± 5.4%) and a maximum F1-Score of 0.90 while retrieving the most similar image from the database. These are promising results for the first known study to demonstrate the feasibility of developing a CBIR algorithm for eardrum images.

We investigated a novel approach, called OtoPair (Camalan et al. 2021), which uses paired eardrum images together rather than deciding on a single eardrum image to detect the eardrum abnormality. OtoPair increases the accuracy from 78.7% (±0.1%) to 85.8% (±0.2%) on a three-fold cross-validation scheme. This preliminary work demonstrated the feasibility of using a pair of eardrum images instead of single images to enhance the diagnostic accuracy. Finally, we proposed a digital otoscopy video summarization and automated diagnostic label assignment model (Binol et al. 2021) that benefits from the synergy of deep learning and natural language processing. Our main motivation was to obtain the key visual features of eardrum diseases from their short descriptive reports provided by an ENT physician (author ACM). The proposed model provided an overall F1-Score of 90.2% to predict the test instance’s diagnostic label of the test instance. To the best of our knowledge, this was the first study to utilize textual information in computerized ear diagnostics. A summary of the literature of CAIA approaches on the ear diseases is presented in Table 1.

**Table 1.** Summary of the literature

| Authors | Purpose | Methods | Database size | Performance |
| --- | --- | --- | --- | --- |
| Mironica *et al.* (Mironică, Vertan, and Gheorghe 2011) | Otitis media detection | Artificial neural networks with global features such as color descriptors | 186 single otoscopic images | Detection accuracy: 73.1% |
| Kuruvilla *et al.* (Kuruvilla et al. 2013) | Otitis media classification | Decision trees with hand-crafted features | 181 single otoscopic images | Classification accuracy: 89.9% |
| Shie *et al.* (Shie et al. 2014) | Otitis media detection | Adaboost-based classifier with texture-based features | 865 single otoscopic images | Classification accuracy: 88.06% |
| Myburgh *et al.* (Myburgh et al. 2016) | Otitis media classification | Decision tree with pre-defined clinical and visual features such as Malleus bone visibility and eardrum color | 489 single otoscopic images | Classification accuracy: 80.6% |
| Lee *et al.* (Lee, Choi, and Chung 2019) | Binary classification of normal and perforated eardrum images | CNN | 1,338 images | Perforation detection accuracy: 91.0% |
| Tran *et al.* (Tran et al. 2018) | Binary classification of AOM and effusion | Multitask joint sparse representation with image features such as color, and shape | 1,230 otoscopic images | Classification accuracy: 91.41% |
| Senaras *et al.* (Senaras et al. 2017) | Eardrum abnormality detection | Fuzzy Stacked Generalization with clinically- motivated<br>hand-crafted features | 247 otoscopic images | Classification accuracy: 84.6% |
| Senaras *et al.* (Senaras et al. 2018) | Eardrum abnormality detection | Ensemble of two deep learning method | 409 otoscopic images | Classification accuracy: 84.4% |
| Binol *et al.* (Binol, Moberly, et al. 2020a) | Eardrum abnormality detection | Ensemble of otoscopy and tympanometry | 73 otoscopic images | Classification accuracy: 84.9% |
| Kasher *et al.* (Kasher 2018; Senaras et al. 2017) | Otitis media classification | CNN with clinically motivated features | 108 otoscopic images | Classification accuracy: 82.2% |
| Binol *et al.* (Binol et al. 2021) | Multiclass classification | NLP using ENT reports | 108 otoscopic images | Classification performance: 90.2% (F1-Score) |
| Camalan *et al.* (Camalan et al. 2020) | Content-based image retrieval | Similarity metrics with deep features | 454 otoscopic images | Retrieval performance: 90.0% (F1-Score) |
| Camalan *et al.* (Camalan et al. 2021) | Eardrum abnormality detection | Paired information of eardrum images with hand-crafted features | 150-pair (300-single) of otoscopic images | Average accuracy of 3-Fold cross-validation: 83.5% ($\pm 0.2\%$) |

All of these prior studies, except our recent study (Binol et al. 2021), require a single image rather than raw videos to analyze eardrum abnormalities (Kasher 2018). This single image needs to be carefully selected manually. Yet, manually selecting a representative frame (also known as the keyframe) from even a few seconds of eardrum videos is extremely time-consuming, requires expertise, and is subject to high inter- and intra-reader variability (Jeffay and Zhang 2001; Han, Hamm, and Sim 2011; Gygli et al. 2014; Binol et al. 2021; Moberly et al. 2018). The complex topology of the eardrum requires multiple images at varying depths of focus to capture the eardrum properly. For this reason, even if a single best frame could be selected from the video, it might not contain the entire view of the eardrum. Therefore, a clear and comprehensive composite image generated automatically from otoscope video clips would be helpful for accurate and automated diagnosis of eardrum abnormalities. Thus, we developed a method called *Stitch*, which creates a composite image by stitching all the frames from the same video data.

After realizing that some of the video frames could be of poor quality (e.g., those that are out-of-focus or those that have noticeable glare), we further enhanced this method and called it *SelectStitch* (Binol, Moberly, et al. 2020b). Five ENT physicians reviewed 78 SelectStitch, Stitch, and full otoscopy videos in a follow-up clinician reader study. They provided equivalently accurate diagnoses when reviewing SelectStitch composite images (60.8% correct) versus full video clips (62.8% correct); however, the performance with Stitch was worse (33.1% correct) (Binol, Niazi, Essig, et al. 2020). These findings suggest that SelectStitch provides composite images equally as diagnostic as viewing the entire otoscopy video clip. However, it was noteworthy that ENT diagnostic accuracy rates were still relatively poor, providing continued evidence for additional computerized method development.

Analyzing video sequences as opposed to a single frame or a composite image presents unique challenges. For example, some of the video sequence frames do not contain useful information because of the poor quality of the images. For this reason, in this study, we determine informative frames after reducing the blurry and glare-affected frames from the video sequence. The new OtoXNet approach is different from our previous approach (Binol, Moberly, et al. 2020b) in that we now generate two composite images out of those frames with a high potential of containing pathology in the eardrum. Therefore, we re-cast the video sequence analysis as the analysis of multiple composite images, which transforms the classification problem into a standard image classification problem. Since the proposed method does not require frame-level labels for supervised training, it can be considered as a weakly-annotated (Prest et al. 2012) classification scheme. As a result, in this paper, we compare three different schemes: 1) transfer learning utilizing a pre-trained deep convolutional neural network (DCNN) on two composite images per video, 2) single composite image case, and 3) a human-selected keyframe picked from each video to represent the corresponding abnormality. We will compare each scheme’s performance in terms of accuracy, sensitivity, specificity, precision, and F1-Score. We used ResNet-101 architecture (He et al. 2016) pre-trained on ImageNet (Deng et al. 2009) as it showed competitive performance on medical imaging applications (Zhang et al. 2020). Figure 1 shows the overall process of the proposed method for the otoscope video classification task.

**Fig. 1.**
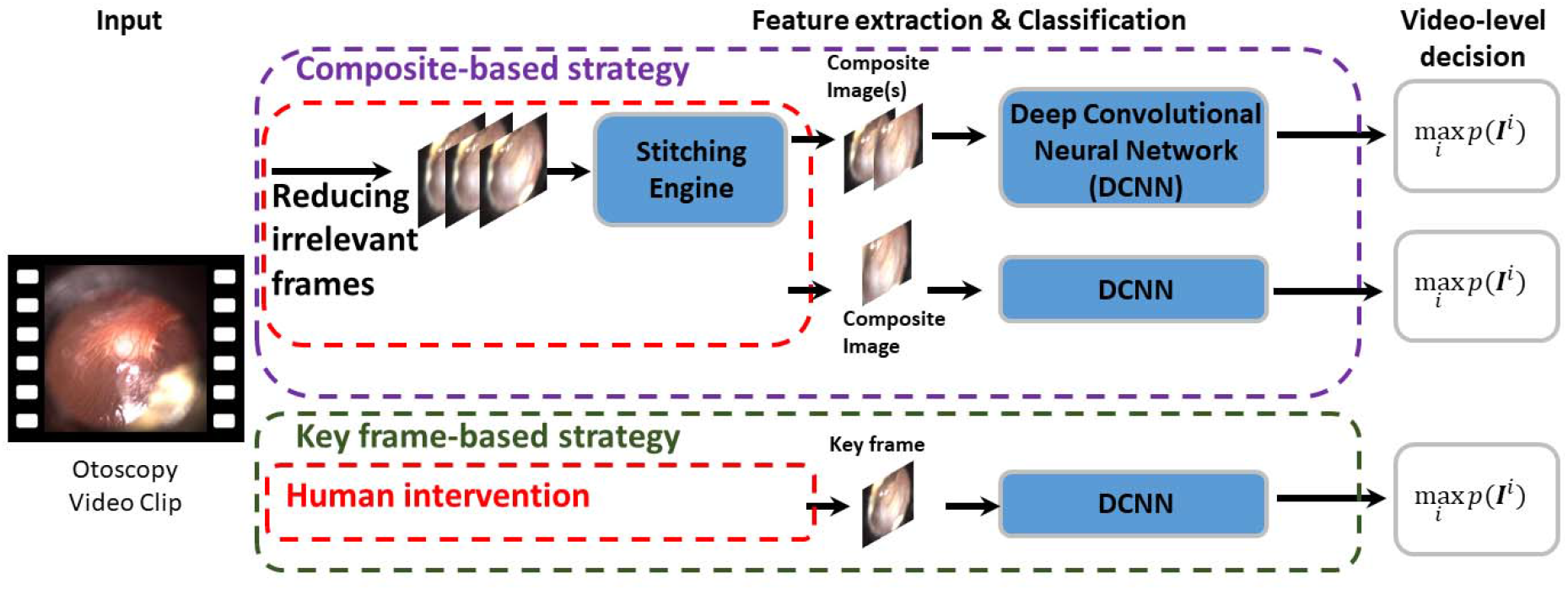
An overview of the otoscope video analysis involved for the video classification. Input is an otoscopy video clip of a patient. The feature vector’s size for an image extracted by DCNN depends on the exact nature of the network used and the layer picked for feature extraction in that network. For example, it equals 2048 for ResNet-101 if features were extracted at the end of the network. The video-level decision is established through the observation of the per-category likelihood ratio score for each composite image: *p*(*l*^*i*^) indicates the likelihood calculated as the classification score where *l^i^* is the *i*^*th*^ composite image, *i* = 1,2 for the original dataset, of the test video.

This paper makes the following novel contributions:

- Unlike previous works that use human-selected keyframes, this is the first work to utilize the otoscope videos directly. As the diagnostic label assigned by the ear experts belongs to the video and not necessarily to each frame, a weakly-annotated classification scheme is employed. We expect that this study will promote the diagnosis of eardrum abnormalities utilizing otoscope videos (Section 2.1);
- A blurriness index is defined to properly pick in focus frames (Equation 3);
- The proposed method determines the meaningful region of an eardrum using α-shapes (Section 2.3);
- We now generate two composite images out of video frames automatically (Sections 2.4, 2.5).

In the following sections, we describe how OtoXNet works and assess its performance. First, Section 2 gives the details about the data and describes the proposed OtoXNet method. Experimental results are introduced in Section 3. We discuss the results and conclude the paper in Sections 4 and 5, respectively.

## 2. Materials and Methods

### 2.1 Materials

Three ENT physicians (authors ACM, GE, and CE) from The Ohio State University (OSU) and Nationwide Children’s Hospital (NCH) in Columbus, Ohio, participated in this study. They have created a database of high-resolution digital adult and pediatric otoscope videos with corresponding diagnostic labels with Institutional Review Board (IRB) approval and patient consent. The collected otoscope videos contained no identifying protected health information ensuring the inability to link them to any particular patient.

A high-definition (HD) video otoscope (JEDMED Horus+ HD Video Otoscope, St. Louis, MO) was utilized to acquire the videos. This tool is an all-in-one handheld digital otoscope with a liquid crystal display (LCD) viewfinder that the user controls to assist in gathering the video. It offers an on-screen display of image sequences for instantaneous review. After each examination, videos were delivered for storage on a secure server right away (Moberly et al. 2018). The video frames are initially of size 1440 by 1080 pixels and are recorded in an MPEG 4 video container file format. The database comprises almost all major eardrum diagnostic categories such as normal, AOM, effusion, perforated eardrum, myringitis, tympanosclerosis, retracted eardrum, cholesteatoma, and eardrum with ventilation tube. We have eliminated some of the categories, e.g., AOM, which did not have sufficient samples necessary to train our algorithms properly. Four categories of otoscopy examination videos, i.e., effusion, eardrum perforation, tympanosclerosis, and normal (healthy), have been included for the qualitative experiments in this study. An example frame for each category from our dataset is depicted in Figure 2. The descriptions of our dataset are given in Table 2.

**Table 2.** Specifications of our otoscope video dataset. All the images were of size 1,440 × 1,080 pixels.

| **Diagnostic Categories** | **Number of Videos** | **Number of Frames (in total)** |
| --- | --- | --- |
| Effusion | 114 | 43,417 |
| Normal | 169 | 35,505 |
| Perforation | 56 | 19,732 |
| Tympanosclerosis | 55 | 13,063 |
| Total | 394 | 111,717 |

**Fig. 2.**
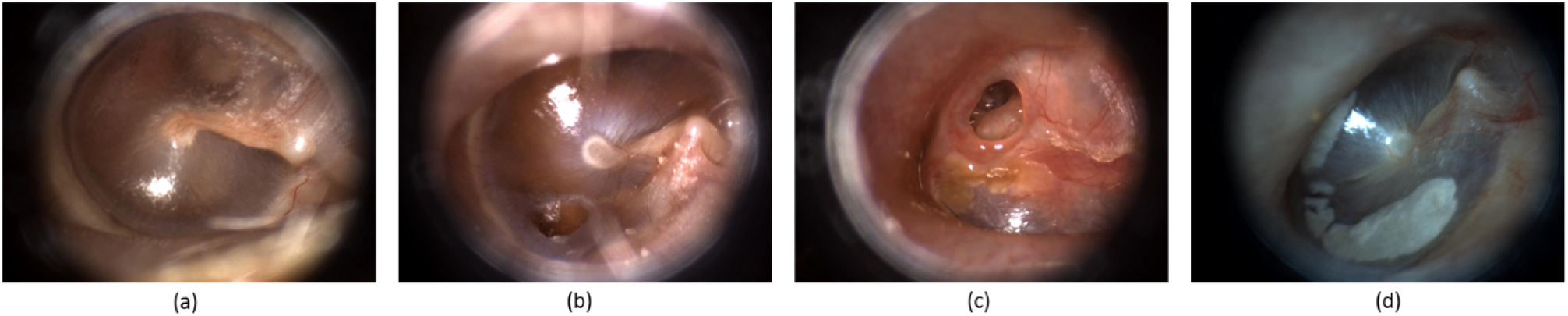
Examples of eardrum conditions observed in our dataset: (a) normal, (b) otitis media with effusion, (c) eardrum perforation, and (d) tympanosclerosis.

### 2.2 Gaussian Filtering

An otoscope often results in videos where frames suffer from motion blur. This occurs when the shutter speed of the video camera is slower than the physical movement of the video camera. In the presence of motion blur, identification of visual features in the image becomes problematic even for a human. In this subsection, we present an image quality assessment technique for blurred images, which will determine the quality of an image based on the amount of blurriness in the image.

Image smoothing is a classical technique for digital image processing and can be accomplished by employing different types of filters. We utilized the most common one, i.e., Gaussian filter, in this study. The edge (the boundary between two regions of different intensity level) detection techniques frequently choose the 2D Gaussian filters (Khorbotly and Hassan 2011). The significant variations can be boosted through subtraction of a Gaussian image from the corresponding original image. Let *I*(*x,y*) and *I*_*G*_(*x,y*) be the original frame and the smoothed version of it by Gaussian filter, respectively. The difference image *I*_*D*_ can be represented by

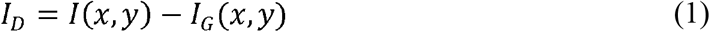

where the Gaussian image can be obtained in the spatial domain via the convolution process;. *I*_*G*_ (*x,y*) = *I* (*x,y*) ⊛ *G* (*x,y*) Here, *G* (*x,y*) is a 2D Gaussian function. In this study, a Gaussian smoothing filter with σ = 5 is applied using a 31 × 31 kernel. σ is determined by an observational study on a small dataset. We examined the 40 frames in total from different videos and inspected the sharp and blurry regions in each frame. The Gaussian filter’s final size is chosen as a larger value than the average size of the meaningful and sharp area of frames. Briefly, we utilized unsharp masking algorithm without getting the sharpened image at the end (Deng 2010). We only extracted the image details instead.

Figure 3 shows the steps followed by the focus area candidate detection technique for two different frames, which have different blur effects, of the same otoscope video clip. It is important to mention that a grayscale version of the input frame is used in this process. The candidate areas to highlight the sharp regions are obtained using the smoothed frame’s subtraction from the original grayscaled one, then calculating the absolute value of the difference image since it has negative values, and finally, an experimentally determined threshold value of ten.

**Fig. 3.**
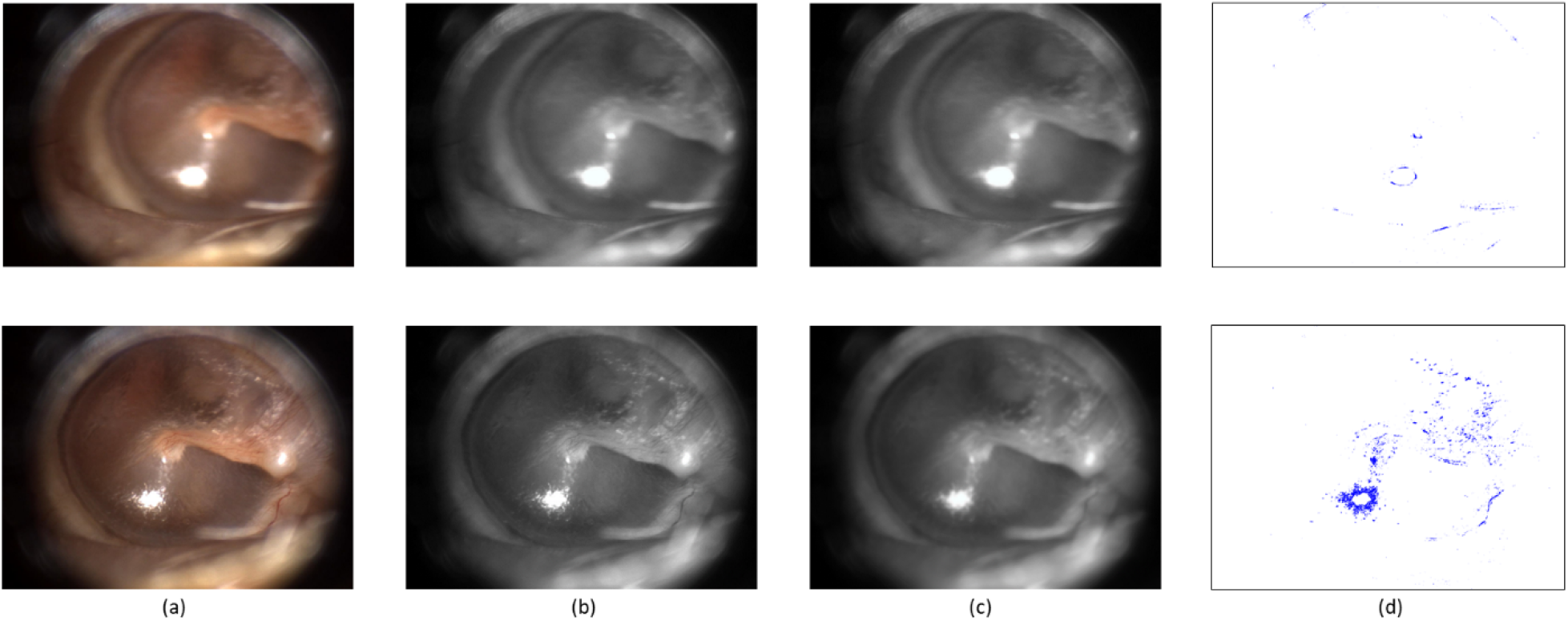
Applying Gaussian filtering on different frames of the same video. Please note that the top frame is blurrier than the bottom frame. For each row: (a) Original frame; (b) Gray level version; (c) Frame smoothed by Gaussian kernel; (d) Details of the original image by subtracting (c) from (a) and thresholding the output.

### 2.3 Determining the Meaningful Region of an Eardrum using α-shapes

Once we obtained the point cloud of “sharps” (Fig. 3(d)) as a discrete representation of the eardrum surface, we need to find the shape of the point cloud so that we can build a measurement of the usability of a frame for CAIA. We modeled the shape of the point cloud using a concave hull. Unlike convex hull, concave hulls are not unique, i.e., there exists no unique solution to computing convacity(Jiang, Lou, and Scott 2011). To establish a method for obtaining usable regions in an eardrum image, we employed the idea of α-shapes (Edelsbrunner, Kirkpatrick, and Seidel 1983). Determining the meaningful region of an eardrum comprises deriving the 2D surfaces from the initial set of points. The α-shape characterizes the perceptive view (visually intuitive) of “pattern” for a set of points in the given scope. For a given real parameter α and a set *S* which includes points, the shape of *S* obtained by α-shape is a polytope, i.e., the generalization to any dimension of a two-dimensional polygon and a three-dimensional polyhedron. Figure 4 illustrates how α–shape helps to transit from sharp points, as determined after the difference imaging method described in Section 2.2 to shapes to determine both focus areas and exclude redundant regions (e.g., glare in this example).

**Fig. 4.**
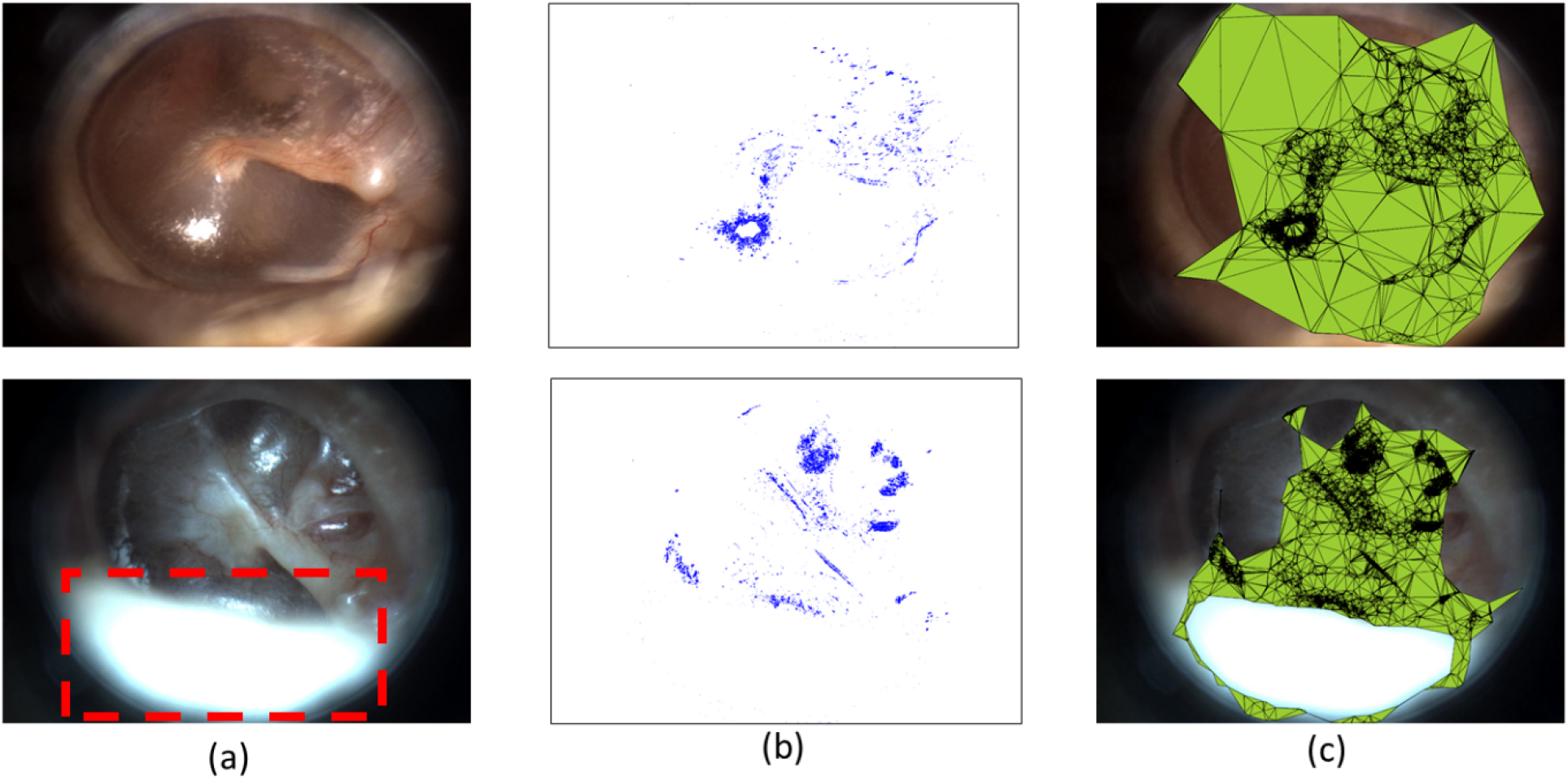
The transition from candidate areas to shapes using α–shape. For each row: (a) Original frame; (b) Output of subtraction and thresholding described in Section 2.2 (Fig. 3(d)); (c) Output of α–shape represents the shape of focus region and excludes redundant region (bottom of Fig. 4(c)). Please notice that the top original frame was also given at the bottom of Fig. 3. A blurry index at this point can be defined as the number of sharp points (pixels) (see Fig. 3(d) or Fig. 4(b)) divided by the area of the whole α–shape region (the green region in Fig. 4(c)). The frame with a glare (the red-dashed rectangle given at the bottom of Fig. 4(a)) is also detected by computing the glare area compared to the meaningful area.

A *blurriness index* can be defined as the number of sharp points (exact area of the sharp areas) (see Fig. 3(d) or Fig. 4(b)) divided by the area of the whole α–shape region (see Fig. 4(c)). This process can also be cast as finding the density of sharp areas. For a given image *I*, let *S* be a set of sharp points on that image, we denote the density of points with *d*(*S*), that is

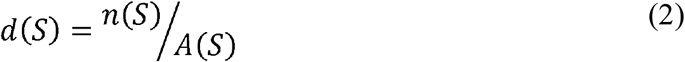

where *n*(*S*) is the number of points (pixels) and *A*(*S*) is the area of the shape obtained by the α-shape. Then *d*(*S*) values were rescaled in a range of 0 to 1 by using min-max scaling among each frame in the same video. Thus, for a video frame *I*, the *blurriness index* of that frame (*b*(*I*)) is given as:

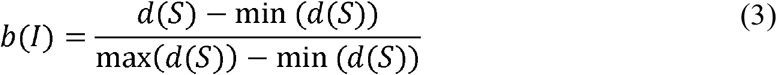

where min(*d*(*S*)) and max(*d*(*S*)) denote the minimum and the maximum of the unscaled variables in the samples. An observational study from the small dataset revealed that the top twenty percent (between 0.8 and 1) and the following thirty percent interval (between 0.5 and 0.8) of the frames having higher *b*(*I*) values are reasonably good to generate multiple composite images. For the single composite case, only the first composite image, i.e., the image generated by the top twenty percent of the frames, was adopted.

Glare is another substantial problem encountered in otoscope videos (note the red-dashed rectangle given at the bottom of Fig. 4(a)). We reduced the video frames with a “remarkable” glare by computing the glare area compared to the meaningful area determined by α–shape. We employed an HSV color space-based white area detection outside of the shape. If the proportion of the area of the white region, which implies glare to the area of the shape, is greater than ten percent, we ignored that frame and did not use it in the image stitching phase.

### 2.4 Composite Image Generation

A set of image stitching techniques in the computer vision field offers different approaches for generating composite images under two main methods, i.e., direct approach and feature-oriented method. In the direct approach, the camera parameters and transformation matrix are calculated by minimizing an error function based on the intensity difference of the overlapped region (Wei et al. 2019). The direct approach might yield an accurate registration, but it fails in stitching tasks when scaling or illumination changes or when there is a considerable amount of noise. Feature-oriented approaches detect the features, for example, edges, points, corners, lines, or other shapes, and calculates the relationship between these features among images. A feature-oriented approach manages locality around detected features to indicate that feature as a key feature known as descriptor (Bouguet 2001). All key features in a pair of images are then compared with that of every feature in another image by adopting the descriptors (Samsudin et al. 2013). For example, OpenCV utilizes an affine transform estimation between neighboring fields using Speeded Up Robust Features (SURF) (Bay, Tuytelaars, and Van Gool 2006) key points matching. These transforms describe the translation, rotation, and skew of each peripheral field relative to the central field. Finally, a composite image is generated using the estimated transforms, and overlapping regions are blended linearly.

For the image stitching algorithm, we used a freeware (for non-commercial use) software package called Image Composite Editor (ICE) 2.0 (Microsoft), developed by the Microsoft Research Computational Photography Group. Our main goal here is to summarize the videos by generating seamless composite images out of their frames. It is worth mentioning that Microsoft ICE does not have the proficiency of stitching images from totally different scenes, e.g. when the initial frames include potential glare and ear canal, and the following frames consist of the eardrum. Accordingly, as proposed in Section 2.3, reducing eardrum-irrelevant frames was needed as a preprocessing step to utilize this tool. Our solution provides a beneficial mechanism by replacing human intervention, e.g., manual keyframe selection.

### 2.5 Transfer Learning with DCNNs

Transfer Learning (TL) (Pan and Yang 2009) is a technique that enables storing knowledge achieved during learning a source problem (*T*_*S*_), and employing that knowledge to learn a target task (*T*_*T*_) efficiently (Lu et al. 2015). In DCCNs, this is accomplished by accepting the weights of a model trained for an initial problem to initialize another model set to be trained for the target problem (Binol, Plotner, et al. 2020; Niazi et al. 2018; Binol, Niazi, Plotner, et al. 2020). Although the similarity between the source and target tasks and the domains impacts the performance of TL, it is stated that applying TL is still beneficial compared to starting the weights from scratch (Yosinski et al. 2014). In this study, we used the DCNN network that was developed for the ImageNet LSVRC-2015 (Deng et al. 2009) contest, in which 1.2 million images were classified into 1000 classes. The ResNet-101 architecture won the ILSVRC-2015 classification, detection, and localization (with a top-1 and top-5 error rate of 19.87% and 4.60%, respectively) (He et al. 2016) and COCO-2015 detection and segmentation competitions (Lin et al. 2014). We used the MATLAB 2020a using the Deep Learning Toolbox in a high-performance computer (HPC) with 128 GB RAM and 16 GB NVIDIA Tesla P100 PCI-E GPU to train the DCNNs.

The ResNet-101 has 101 layers deep with rectified linear units, max-pooling layers, and residual connections. ImageNet of ResNet-101 was trained with an input ROI size of 224 × 224 pixels RGB channel. We applied fine-tuning instead of “vanilla” transfer learning which is based on taking the pre-trained network weights learned from a larger benchmark dataset, ImageNet in this case, and re-train the fully-connected layers on the target data. We froze the weights of the first 291 (out of 347 in the MATLAB R2020a environment) layers of ResNet-101 to transmit the generalized features from these layers for transfer learning while re-training the remaining convolution layers and the fully connected layers for the task at hand. We applied additional dropout methods in a fully connected layer to reduce overfitting. For our eardrum disease classification task in otoscopy, one additional fully connected layer (*fc*_4_) was inserted instead of *fc*_1000_ to the original structure to drop down the 1000-node fully connected layer to a 4-node soft-max classifier.

All the fully connected layer weights and biases were randomly initialized for retraining. We also changed the cross-entropy loss function to weighted cross-entropy loss before fine-tuning. A weighted classification layer computes the weighted cross-entropy loss for classification problems. Weighted cross-entropy is an error measure between two continuous random variables. For prediction scores *y* and training targets *t*, the weighted cross-entropy loss between *y* and *t* is given by

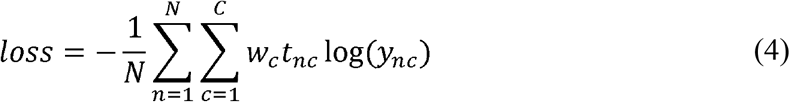

where *N* is the number of observations, *C* is the number of classes, and *w* is a vector of weights for each class.

## 3. Experimental Setup and Results

### 3.1 Experimental setting

To reduce the effects of overfitting and enhance the training stability, we took advantage of data augmentation (Chen and Lin 2014): each image was rotated from -15 to 15 degrees in 3 degrees intervals and each output image was scaled in [0.75, 0.8, 0.85, 0.9, 0.95, 1, 1.05, 1.1, 1.2]. Hence, we produced 99 more images for each original sample. Moreover, we also applied on-the-fly augmentation by randomly flipping them on x-direction and translated in x and y directions within the range of [-50, 50]. It is important to note that the classification framework during training was not exposed to the validation/test images nor their augmented versions. Although each ear might have a different abnormality from its pair (Camalan et al. 2021), we put the same patient’s video recordings into the same group of validation. To address the problem of class imbalance in our study (see Table 2), we practiced random oversampling, which involves randomly selecting examples from the minority class (perforation and tympanometry in our case), with replacement, and adding them to the training dataset (Yap et al. 2014).

In total, we train for 10 epochs using Adam optimizer (Kingma and Ba 2014), starting with a learning rate of 1 × 10^−5^ and dropping it by a factor of 0.8 after every two epochs. We also conducted early stopping by reserving ten percent of the training data as a validation set. We use a batch size of 64 on a single NVIDIA Tesla P100 PCI-E GPU. Using an 8-fold cross-validation scheme, three different eardrum identification methods were investigated. A total of 394 otoscope videos were consumed (see Table 2). First, a pair of two composite images were generated (see Section 2.4). A pre-trained ResNet-101 was applied to the composite images for a deep learning-based system (see Section 2.5). Although we experimented with longer and shorter training times, additional training did not lead to a noticeable improvement. We also conducted additional experiments with other cross-validation schedules such as 24-fold and leave-one-patient-out, but they were not presented here as they gave no additional improvements.

The video-level decision is established through the observation of the per-category likelihood ratio score for each test image:

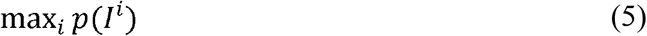

where *I*^*i*^ is the *i*^th^ image, *i* = 1, …,100 for keyframe and single composite image cases, and *i* = 1, …,200 at max for multiple composite case, of the test video and *p*(*I*^*i*^) indicates the likelihood calculated as the classification score.

For all of these experiments, image pre-processing is kept to a minimum after the composite image generation step (Section 2.4). Composite images are resized to 224 × 224 and the channel means are normalized for ResNet-101. To investigate the effectiveness of the DCNN-based methods, we presented the performance metrics in terms of Sensitivity (*TP*/(*TP* + *FN*)), Specificity (*TN*/(*TN* + *FP*)), Precision (*TP*/(*TP* + *FP*)), F1-Score 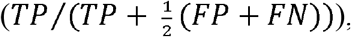, and accuracy (*TP* + *TN*)/(*TP*+*TN* + *FP*+*FN*)where *TP* is the number of positive video instances correctly classified, *FN* is the number of positive instances misclassified, *FP* is the number of negative instances incorrectly classified, and *TN* is the number of negative instances correctly classified.

### 3.2 Evaluation – Experimental Results

To assess the impact of the data augmentation in the performance for OtoXNet, we present the performance values for the manual keyframe selection, the single, and the multiple composite images cases. Data augmentation was more accurate and enhanced the training stability in both cases. As we observed from these experiments, the proposed method of using multiple composite images results in superior overall performance compared to using both a single composite image and a keyframe. The original dataset’s performance values indicate that using multiple composites helps increase the sensitivity by 5.8%, the specificity by 0.5%, the precision by 1.8%, and the F1-Score by 3.7% compared to the single composite image. These values respectively are 6.6%, 2.2%, 9.9%, and 9.8% higher compared to those of the keyframes. With the augmented data, the multiple composites improved the sensitivity by +5.4%, the specificity by +1.6%, the precision by +4.7%, and the F1-Score by +5.3%, compared to that using single composite images, and the sensitivity by +3.4%, the specificity by +1.1%, the precision by +3.0%, and the F1-Score by +3.3% in the comparison with the keyframes. An interesting observation was that the data augmentation for the keyframe case showed a larger increase (+14.5% in F1-Score) in classification performance in contrast to utilizing it for the single composite images (+6.4% in F1-Score). These results are summarized in Table 3. Please note that the overall scores show the final values computed on the entire dataset.

**Table 3.**
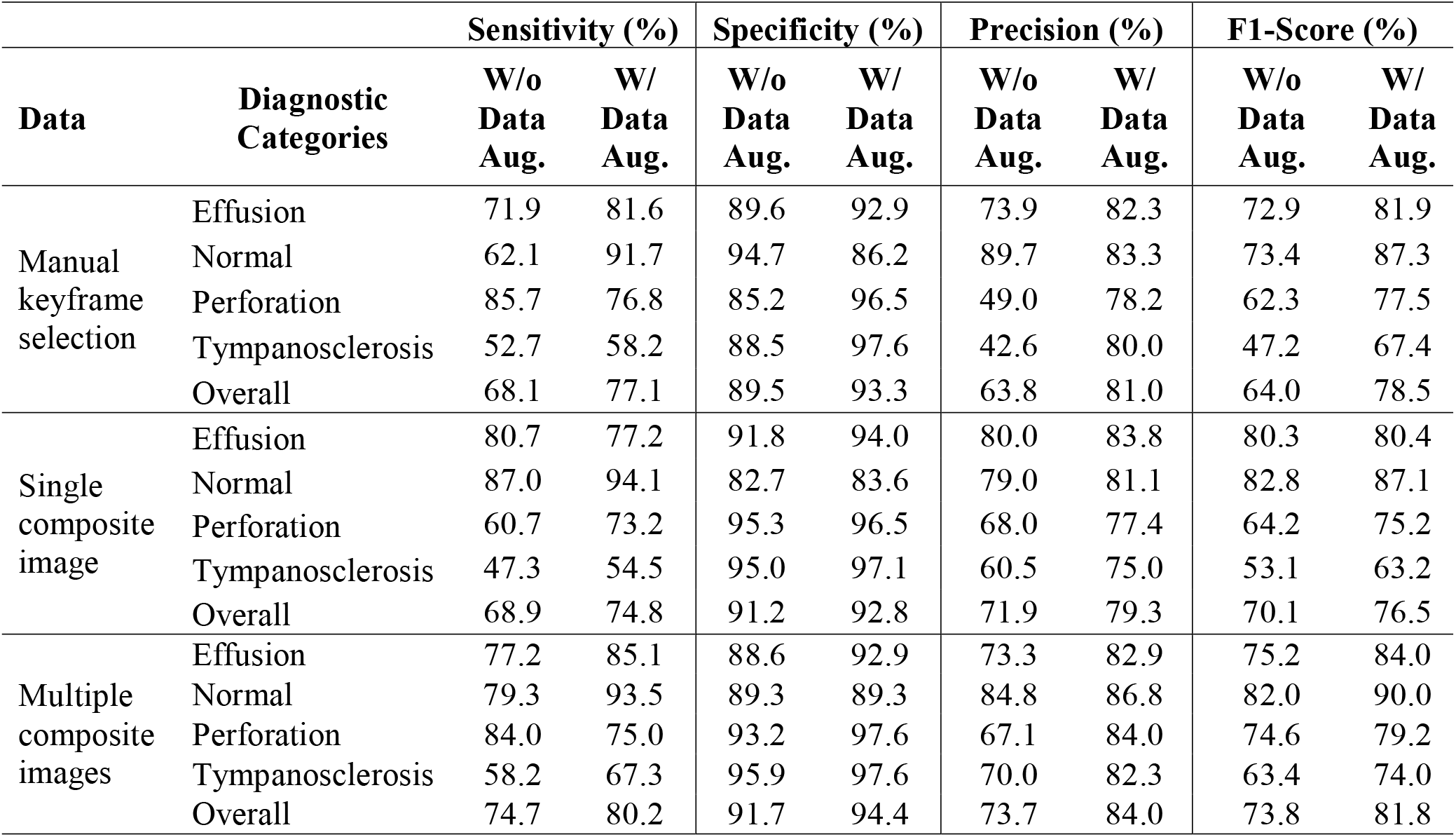
Performance Comparison of OtoXNet (Single and Multiple Composite Images) with keyframes in case of with (w/) and without (w/o) Data Augmentation. Overall scores are computed over the entire dataset.

#### Effect of oversampling and class-weighted loss function

To investigate the effectiveness of the oversampling and class-weighted loss function on the individual diagnostic category performances, we conducted experiments without employing those techniques. Fig. 5 shows that applying the random oversampling and class-weighted loss function process has improved the performance of the minority categories (especially tympanosclerosis) in the case of multiple composite images.

**Fig. 5.**
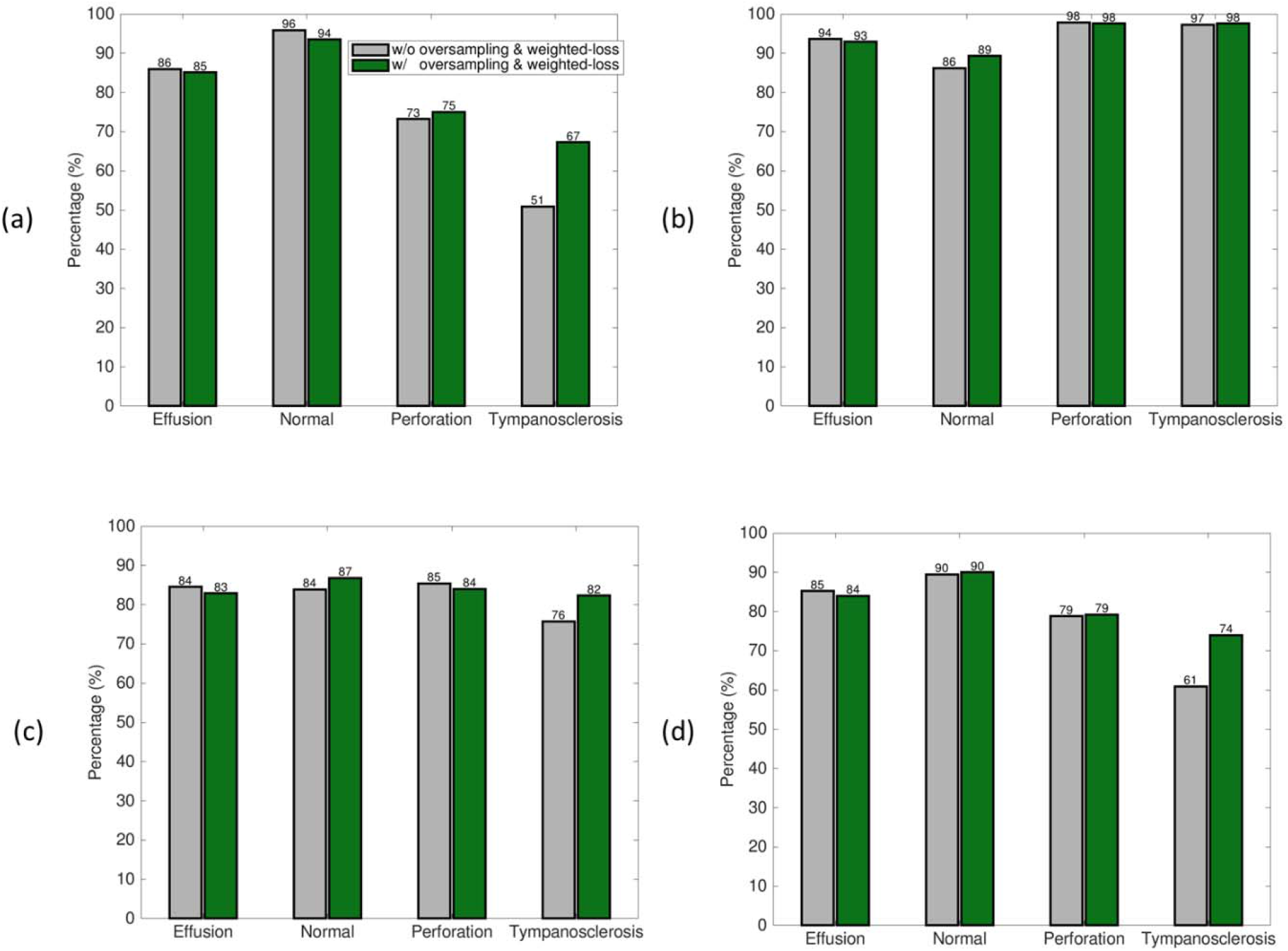
Effect of applying random oversampling and class-weighted cross-entropy loss function with the multiple composite images. (a) Sensitivity, (b) Specificity, (c) Precision, (d) F1-Score.

We also reported the class-weighted average accuracy across cross-validation folds in Fig. 6. While OtoXNet with multiple composite images has demonstrated 84.8% classification accuracy in average (±3.8%) of 8-folds, the keyframes obtained 81.8% with a 5.0% standard deviation. A paired t-test indicated that there is a statistically significant difference between these values (p-value = 1.3 × 10^−2^). The average accuracy for the single composite case was 80.1% with a 4.8% standard deviation and the p-value between single and multiple composite cases (3.4 × 10^−2^) suggests a significant improvement in accuracy when using multiple composite images in the eardrum abnormality classification. On the other hand, the p-value between the folds of keyframe and single composites was 5.49 × 10^−1^. This value indicates that there is a 54.9% possibility that the difference in classification occurred only by chance and is considered statistically not significant.

**Fig. 6.**
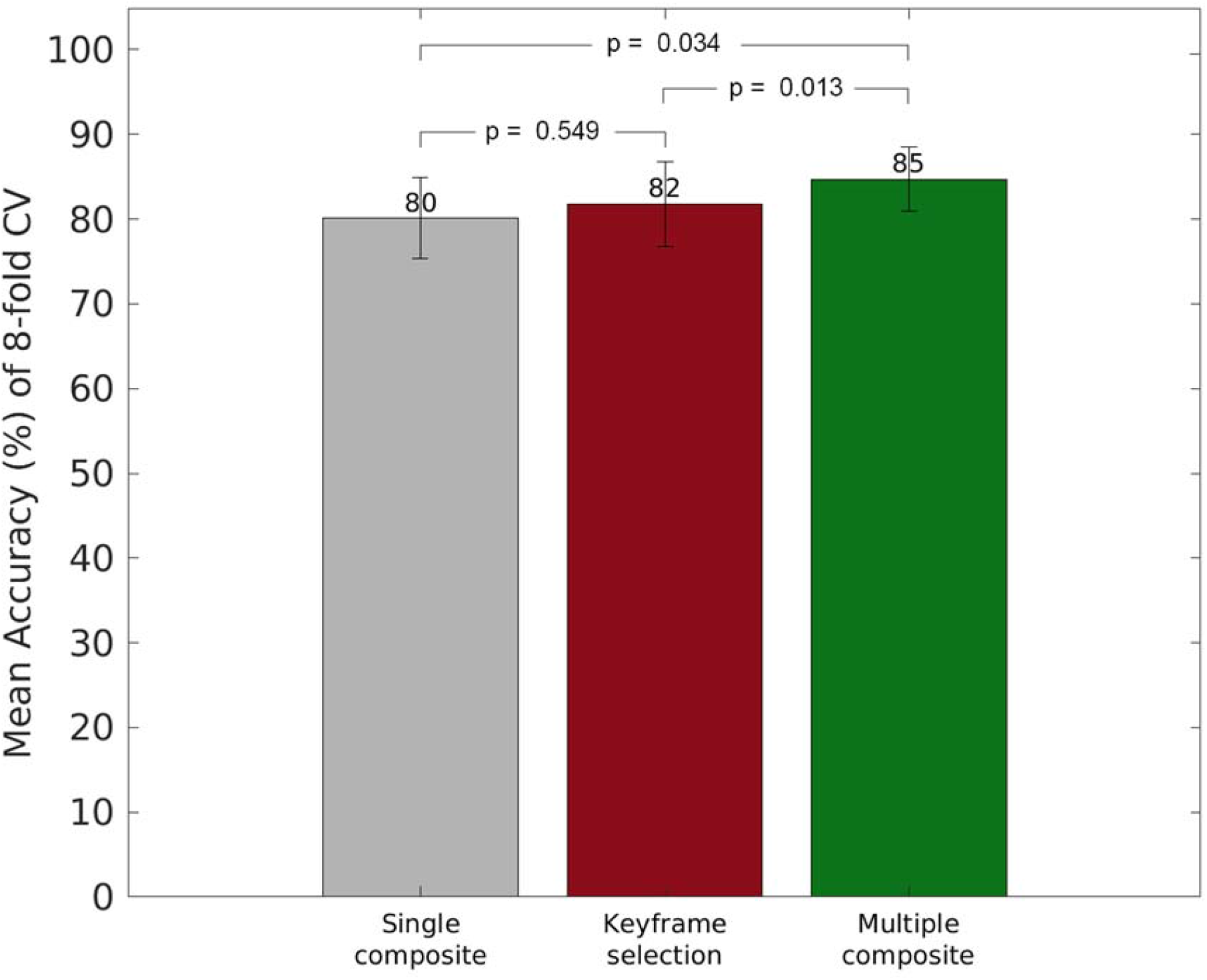
Performance Comparison of OtoXNet (Single and Multiple Composite Images) with keyframes in terms of class-weighted average accuracy across cross-validation folds.

The confusion matrix for the case of OtoXNet (multiple composite images) is given in Table 4. OtoXNet has demonstrated 80.2% of sensitivity (3.1% more than the keyframes), 94.4% of specificity (1.1% more than the keyframes), 84.0% of precision (3% more than the keyframes), and 81.8% of F1-Score (3.3% more than the keyframes) in the overall performance of eardrum video classification task.

**Table 4.** Confusion Matrix for Classification with OtoXNet (Multiple Composite Images)

|  |  | Predicted Label |  |  |  |
| --- | --- | --- | --- | --- | --- |
|  |  | Effusion | Normal | Perforation | Tympanosclerosis |
| True Label | Effusion | 97 | 9 | 4 | 4 |
|  | Normal | 8 | 158 | 2 | 1 |
|  | Perforation | 7 | 4 | 42 | 3 |
|  | Tympanosclerosis | 5 | 11 | 2 | 37 |

## 4. Discussion

In our previous study, we had demonstrated that we could use composite images to classify otoscopy videos. In this study, we expanded that approach by employing multiple (two) composite images out of selected frames. Further, we utilize transfer learning for effective knowledge transfer of generic features for natural RGB non-eardrum images. This work provides a more general pipeline than our previous efforts to diagnose TM as it uses raw videos as an input instead of expert-selected keyframes. A recent study (Alenezi et al. 2021) has shown that video otoscopy increases the ability to make diagnoses, compared to still images.

Classification after applying data augmentation provided superior performance compared to the original dataset, as listed Table 3. A significant improvement was observed particularly for the abnormalities which have relatively limited data, i.e., perforation and tympanosclerosis (please see the F1-Scores of with and without data augmentation for both dataset). This result implied that data augmentation might help to deal with classification over unbalanced datasets. Moreover, the data augmentation for the manual keyframe selection case showed a larger increase in classification performance (particularly in terms of F1-Scores) in contrast to utilizing it for the single composite images (see Table 3).

To assess the effectiveness of video frame elimination using Gaussian filtering and α-shape (see Section 2.2 and Section 2.3), we generated composite images without using these steps, which means stitching all the frames of a video. A pair of composite images for each category with stitching all the frames and only stitching the “good” frames from two different *b*(*I*) intervals determined by OtoXNet are given in Figure 7. Someone can easily infer from Figure 7 that exploring useful features from the stitched images generated by all the frames of a video will be challenging. Moreover, it can be observed that OtoXNet with two composite image case helps to acquire composite images with varying content. The distinguishable differences for effusion, perforation, and tympanosclerosis are demonstrated with the purple-dashed rectangle in the second composite images. Although it may not be seen a significant discrepancy between the images for normal case, we believe that generating multiple composite images still increase the training robustness, exposing the classifier to differing views of the TM.

**Fig. 7.**
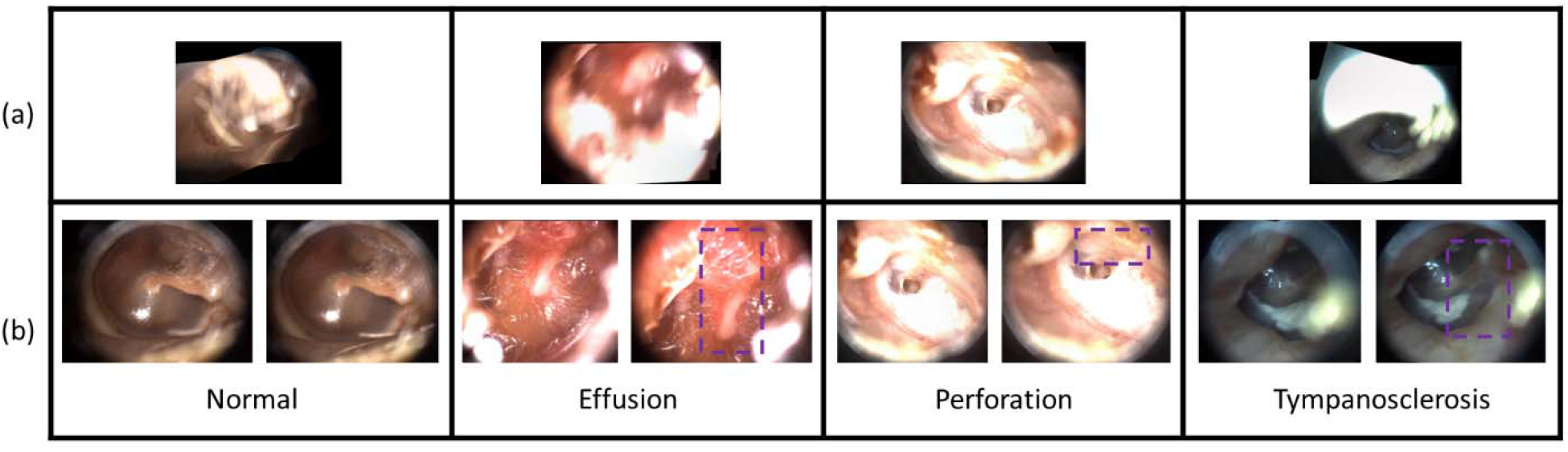
Example composite images of the same video without (a) and with *OtoXNet* for two composite image case (b). For each column, from left to right: normal, effusion, eardrum perforation, and tympanosclerosis. The figure shows the effect of reducing irrelevant (or having low image quality) frames before stitching process using OtoXNet. The first column depicts composite images of a normal eardrum. The images with OtoXNet are obviously clearer as it has minimal glare. The second column shows composite images of an effusion. Again, the images with our method are superiorly clearer and have superior pathology features with less blurriness. Additionally, the second image in OtoXNet focuses on a different region of the frame. The third column illustrates composite images of a perforated eardrum. Here, the images with and without OtoXNet look similar, but the images at the bottom have fewer redundant regions (note the rim of the eardrums). Moreover, the second composite image with OtoXNet has a better view of the top part of the perforation (highlighted with a purple-dashed rectangle). The last column shows composite images of an eardrum with tympanosclerosis (note the white plaque within the eardrum). Again, the bottom images are incomparably clearer and have preferable pathology features with having reasonable glare compared to the top image. As we can see, the second composite image contains a descent view of malleus in this case.

Through transfer learning, a pre-trained DCNN can learn common representations across tasks. In our case, we used transfer learning to construct a diagnostic tool for screening patients with multiple eardrum abnormalities on top of ResNet-101 features. Nevertheless, there are substantial dissimilarities in ImageNet images’ classification, depicting real-life objects and medical images (Raghu et al. 2019). On the other hand, the findings of this study have high significance for eventual clinical impact. The near-85% accuracy of the software compares very favorably with previous findings of 50 to 73% by clinicians (Sorrento and Pichichero 2001). By providing a more objective approach to diagnosing ear diseases through computer-assisted approaches, over-diagnosis and over-treatment should decrease substantially. Moreover, these approaches could be applied in situations where clinicians trained in otoscopy are less accessible, such as low-income countries or rural settings (Myburgh et al. 2016). The videos and images could also be stored using a cloud-based platform, allowing transfer between health institutions for telemedicine purposes. Lastly, the computer-assisted algorithms would be useful in clinician training programs by providing immediate feedback to trainees who are learning to perform otoscopy. In our future studies, we will explore the contribution of OtoXNet to clinicians in their diagnostic workflow.

There are some limitations to this study. First, as we highlighted through Sections 2.2 – 2.4, our method did only focus on the general image quality during the frame selection and stitching process as we believe that the physician should has also viewed the abnormality-responsible area clearly at least in a few video frame period while recording the videos. However, there is a possibility that the automatically selected frames may not contain the abnormality region and further, the stitching engine may ignore those areas during the stitching process. A potential remedy would build a method by learning the clinical and visual features of each category and utilize it on raw video frames without applying stitching. This may require a large amount of data (otoscopy videos) from each category to improve the generalization capability of the detection method. Second, although OtoXNet accomplished competitive performance on the dominant categories, i.e., effusion and normal, and a performance-boosting on the minority ones with data augmentation and applying class-weighted loss function, other categories also need to be studied. Initially, more effective DCNN architectures and/or advanced loss functions, e.g., focal loss, can be applied to increase the classification capability of the methods.

## 5. Conclusions

In this study, we developed a computerized method, called OtoXNet, to make a diagnosis of ear disease from otoscope videos. The proposed method could classify eardrum videos through supervised training and increase identification performance by simultaneously learning features from different tasks (using transfer learning). The videos used in our study were acquired from a hand-held HD video imaging system. OtoXNet shows the strong potential that the eardrum classification task in ENT can be extended to identify other important categories like acute otitis media. On the other hand, as we demonstrated with data augmentation in our study, larger datasets would result in improved performance and more generalizable method sets. The experiments demonstrated that employing multiple composite images in a CAIA system is an effective way to analyze video sequences because they result in a better performance to both human-selected frames and single composite images. We envision that the proposed method has the potential to build a computer-aided diagnosis system to analyze medical videos remotely from a large set of them.

This preliminary work underlines the ongoing need for more objective diagnostic support, and it also suggests the value of developing CAIA approaches that rely on both composite images and raw video clips. For future work, we plan to develop a region detection-based automatic diagnosis method by learning the visual features of each abnormality. By using this approach, we aim to come up with a more interpretable method as it is required to localize the responsible area. For this purpose, we are recruiting more patients to increase the number of cases for the minority categories and also to include AOM in future studies. We are considering that by extending the diagnostic categories of the current study and including region-based features, we will also be able to boost the impact of the OtoMatch by integrating more robust deep features from OtoXNet.

## Data Availability

The data that support the findings of this study are available from the corresponding author, HB, upon reasonable request.

## Acknowledgments

The project described was supported in part by Award R21 DC016972 (PIs: Gurcan, Moberly) from National Institute on Deafness and Other Communication Disorders. The content is solely the responsibility of the authors and does not necessarily represent the official views of the National Institute on Deafness and Other Communication Disorders or the National Institutes of Health. The authors would like to thank Garth Essig, Jay Shah, Theodoros Teknos, Nazhat Taj-Schaal for useful discussions and for the initial curation of the database. The authors would like to thank Ubeydullah Binol for helping us in making Fig. 6 more visually clear.

